# “Trained Immunity” from BCG Vaccination and *Mycobacterium* spp. Exposure may be Linked to Reduced Early Childhood (<5-Year-olds) Brain and CNS Cancer Incidences

**DOI:** 10.1101/2024.10.09.24315142

**Authors:** Samer Singh, Rakesh K. Singh

## Abstract

Globally, with improvements in general hygiene, the incidence of early childhood (0-4Y-olds/<5Y-olds) brain and central nervous system (BCNS) cancers is increasing. Although immunological underpinning is suspected, the identification of protective variables in most BCNS cancer cases remains elusive. Extant hypotheses suggest a role for progressively diminishing exposure to common microbes/pathogens in the rise of childhood cancers in industrialized countries with improved hygiene. Natural exposure to common microbes/pathogens and childhood vaccinations help train the developing immune system of children to respond appropriately to future infections and maintain a healthy immune system. Considering the established role of childhood vaccinations in augmenting immunity, including “trained immunity,” their protective role in pediatric cancers can be surmised. However, a lack of definitive theoretical and practical frameworks to explain conflicting observations has impaired progress. When we analyzed the epidemiological data of European region countries with different childhood vaccination policies but more similar socioeconomic conditions, access to medical services, and genetic makeup as compared to other parts of the world, the coverage of seven major childhood (0-1Y-olds) vaccines was not significantly associated with BCNS cancer incidences in the same cohort of 0-4Y-olds (2020). However, interestingly, prevailing tuberculin immunoreactivity, a surrogate for the existence of heterologous cell-mediated immunity resulting from exposure to *Mycobacterium* spp., including Bacille Calmette-Guérin (BCG) vaccination, for these populations, is found consistently negatively correlated with the BCNS cancer incidence in 0-4Y-olds for countries mandating neonatal BCG vaccination [*r*(24): -0.7226, p-value:<0.0001]. Neonatal immune system priming by BCG and boosting by exposure to environmental *Mycobacterium* spp. appear protective in 0-4Y-olds. Exploration of BCNS cancer incidence and prevailing immune correlates in matched cohorts, along with prospective randomized controlled trials, may be warranted to confirm the impact of childhood vaccinations and boosters (including natural exposure) on early childhood BCNS cancer incidence.

**STATEMENT OF SIGNIFICANCE:** The discovery of preventive variables/interventions for early childhood (<5Y-old) brain and other CNS (BCNS) cancers is highly desirable. The early childhood BCNS cancer incidence rates in countries with differential childhood vaccination coverage and policies display a strong negative correlation with *Mycobacterium* spp. exposure chances/frequency in neonatal BCG-vaccinated children. A potentially protective cause-and-effect relationship with timely *Mycobacterium* spp. exposures (BCGs and/or environmental) seems likely. An assessment of the biological and translational aspects of current observations is warranted.

## INTRODUCTION

Childhood exposure to common microbes, pathogens, and childhood vaccines is supposed to provide appropriate immune training and surveillance capability that generates cross-protective immunity against the number of pathogens to which children are exposed later in life [1-3]. It is associated with better immune function, characterized by reduced allergies to different allergens, and reduced disease occurrence, including cancer [1, 3-9]. Primary brain tumors and associated central nervous system (BCNS) cancers are the most common solid tumors in children (0–14 years old) and adolescents (15-19 years old), accounting for the highest number of solid cancer-related deaths [10,11]. The incidence of childhood BCNS cancers is highest in children less than 5-year-olds (i.e., 0-4Y-olds) that progressively reduces until adolescence before again starting to steadily rise with age during adulthood [10,11]. The BCNS cancers in children are inherently different from those in adults. Those found in children are predominantly malignant gliomas, embryonal tumors, and germ cell tumors, with a minority of non-malignant tumors primarily comprising pituitary tumors [11-13]. A large number of environmental and genetic risk factors have been studied for their association with BCNS cancer incidences [11,12,14] Based on epidemiological studies, exposure to ionizing radiation (increased incidence) and inherited single-gene syndromes (∼4% of childhood cases) are the only validated primary risk factors for BCNS cancer incidence [11,12,14]. Structural birth defects (non-chromosomal) have been associated with approximately 7% of childhood BCNS cancers [11,15,16]. Higher incidence rates have also been observed in the population’s higher socioeconomic position (SEP) sub-groups [17-20]. The “hygiene hypothesis” has been supposed to provide an explanation in the form of a change in SEP-associated unknown risk factor(s) that could alter the exposure of the immune system to common allergens and pathogens, resulting in aberrant immune system development or immune training and an associated risk of childhood cancer [9,11,12,21]. For the majority of early childhood BCNS cancers, the precise nature or identity of risk factors that could be promoting or protecting 0-4Y-old age group children against BCNS cancer incidence remains unknown, while those identified or suspected in a minority of cases lack any interventional value [11,12].

Childhood vaccines play an important role in protecting children from major infections and diseases by augmenting the developing immune system [1]. As the first year of life is important in shaping and training the developing immune system, the recommended immunization schedule [22], starting from birth to the first year of life, generally includes vaccine shots against the following common diseases: tuberculosis (Bacillus Calmette-Guérin, BCG), polio (polio vaccine: POL), Hemophilus influenza B (HIB), hepatitis B (HEPB), pneumococcal illness (pneumococcal conjugate vaccine: PCV), measles infections (measles containing vaccine: MCV), diphtheria, tetanus, and pertussis (DTP). These childhood vaccines are supposed to enhance trained innate immunity and humoral-and cell-mediated immunity in children [1-3,23,24]. Types of these vaccines range from subunit vaccines, heat-inactivated to live-attenuated vaccines, each activating and training a specific combination of the effector arms of the developing immune system [1]. The timing and number of shots vary for each, beginning soon after birth to throughout the entire first year of life, and boosters are often given later in life based on past experiences with vaccines and perceived threats to children’s lives [22]. The first doses of the BCG, HEPB, and OPV vaccines are administered at birth. Two doses of MCV (MCV2), two additional doses of HEPB (HEPB3) and POL (POL3), and three doses of PCV (PCV3), DPT (DPT3), and HIB (HIB3) are administered at different time intervals during the first year of life to generate, strengthen, and sustain immunity against target pathogens, as well as general immunity during the most susceptible period. It may be pertinent to state that the previously recommended booster doses of BCG have been discontinued worldwide due to a lowering of threat perception resulting from a general increase in hygiene and the lowered tuberculosis incidence and associated risk of exposure to the environmental *Mycobacterium tuberculosis* complex. In many industrialized countries, which also happen to have the highest childhood cancer incidence rates, BCG vaccination has been completely discontinued, including the neonatal dose, for the perceived lack of benefit for these countries with their elimination of tuberculosis. The nonspecific positive effect of the BCG vaccine on the survival of children is well recognized [references in 25-27]. It is presumed to be through providing needed immune training to enhance general immune surveillance and strengthening the “trained immunity” of the naïve developing immune system of neonates by functional reprogramming of the immune cells involved in innate immune response [23,28-31] that ultimately seems to cross-protect them against various pathogens and diseases [1,6,24,25-27,32,33].

A number of epidemiological studies around 1970 found that neonatal BCG vaccination was associated with a reduced incidence of leukemia, the most frequent childhood cancer, in up to 6-year-old children [6,34-37]. The proposed mechanisms for the observed reduced cancer incidence in these children have envisaged a role for neonatal BCG vaccinations or infections in priming and training the developing immune system to efficiently eliminate pre-existing pre-cancerous clones or cells—remnants of embryonic development [6,36]. Based on the observed need for a second hit in genetically predisposed models of the disease and the investigation of familial cases of leukemia that are often supposed to get triggered by infections, Greaves hypothesized the need for infections that could lead to overt cancer growth following a second genetic hit [38]. However, many retrospective studies failed to see the protective impact of childhood vaccination and early infections on childhood cancer incidences, with some even suggesting and arguing for an increase in cancer incidence on vaccination [References in 6,39-42]. For a brief account of other theories proposed for childhood leukemia incidence as a result of differential pathogen exposure and immune training, refer to the review by Hauer et al. [43]. Unlike the studies that demonstrate a reduction in the incidence of childhood cancers (leukemia and BCNS cancer) with neonatal BCG vaccination, those witnessing no effect or arguing for an increase in cancer incidences had children vaccinated at times other than the neonatal period [REFs in 37,39-42]. Furthermore, the follow-up cancer incidence periods varied, ranging from beyond early childhood to old age, when the origin and nature of cancers markedly differed from those of early childhood cancers [6,11,12,36,38,43,44]. Some studies even seem to equate a lack of curative potential in age groups other than early childhood with a lack of preventive potential in early childhood. However, due to the lack of any clear-cut epidemiological evidence and any apparent way forward for their validation, the role of any childhood vaccination, including BCG, for childhood cancer prevention has remained highly debated and controversial [6,32,37,39,40,42,44,45].

Based on our observation that the incidence of BCNS cancer in up to 5-year-old children in similar HDI countries worldwide was significantly inversely correlated with tuberculosis incidence rates in these countries, we have suggested that exposure to *Mycobacterium* spp. could be potentially protectively linked to BCNS cancer incidence in young children [45]. In this article, we explore our assertion by presenting an epidemiological analysis of early childhood BCNS cancer incidence in the WHO European region (ER), which has more similar underlying socioeconomic conditions, genetic makeup, comparable health and medical infrastructure access, and disease reporting as compared to the rest of the world. However, there is a diversity in universal vaccination programs/policies that range from never-ever to all eligible for different vaccines, with coverage ranging from 0% to almost 100% [WHO-UNICEF Estimates of National Immunization Coverage (WUENIC): 2022 revision, updated July 2023; https://data.unicef.org/topic/child-health/immunization/] [46] potentially offering a diverse mix of supposed ‘trained immunity’ status from different vaccines and exposure to *Mycobacterium* spp. in this population to identify potential protective determinants. With regard to BCG vaccination policy, the ER region is almost equally divisible in BCG-vaccinating (BCG) and non-vaccinating (No-BCG) countries [The BCG World Atlas, 3rd Edition, http://www.bcgatlas.org/index.php] [47]. The existence of populations in the European region with 50 different levels of prevailing tuberculin immunoreactivity (TI), also referred to as ‘latent tuberculosis infection’ (LTBI) [48-51] or TST positivity—a surrogate measure of existing ‘trained immunity’ and cell-mediated immunity-boosting potential from exposure to *Mycobacterium* spp. (BCG or/and environmental) [51-55], offers an excellent opportunity to examine our assertions.

## RESULTS

### Early childhood vaccinations and incidence of BCNS cancer

When the ER-countries, with comparable confounders, contributing to 15% of global BCNS cancer incidences in 0-4Y-olds during 2020 (GLOBOCAN 2020; https://gco.iarc.fr/today) [56] and displaying almost two times higher age-standardized incidence rate (ASR) as compared to the rest of the world (2.4 vs. 1.3 ASR per 100,000; range: 0-5.4 ASR), are examined to identify potential impact of different early childhood vaccinations, the coverage of BCG or any other test vaccines (*i.e.*, DTP3, MCV2, PCV3, POL3, HEPB3, HIB3), in the same cohort of children, i.e., during 2016-2020 in 1-year-olds (WUENIC) [46], do not significantly correlate with the BCNS cancer incidence rates (Table 1)[22,46]. This apparent dissociation observed in the population-level data seems to support the contention that childhood vaccinations do not play any significant role in protecting against childhood cancers [reviewed in 39, 40], contradicting previous observations made in the 1970s on 0-to 6-year-old children-associated data analysis [34-36]. However, among all childhood test vaccines assessed, the negative correlation, although non-significant and very weak, seemed strongest for the BCG coverage of countries reporting non-zero BCNS cancer incidence in 0-4Y-olds [*r*(47): -0.1667, *p*-value: 0.2628](Table 1). Countries with a small population size and reporting zero incidence for 2020 (i.e., Iceland, Malta, and Luxembourg) were excluded from the analysis. Since BCG-vaccination-elicited immune responses in children are short-lived and wane away within years in the absence of boosting from rechallenge with BCG vaccine or environmental *Mycobacterium* spp. exposure [53,57,58], the observed lack of a correlation for BCG coverage would be expected in the current populations with higher hygiene standards and lower chances of boosting from re-exposure to *Mycobacterium* spp., even if BCG vaccination could have been protective.

**Table 1.**
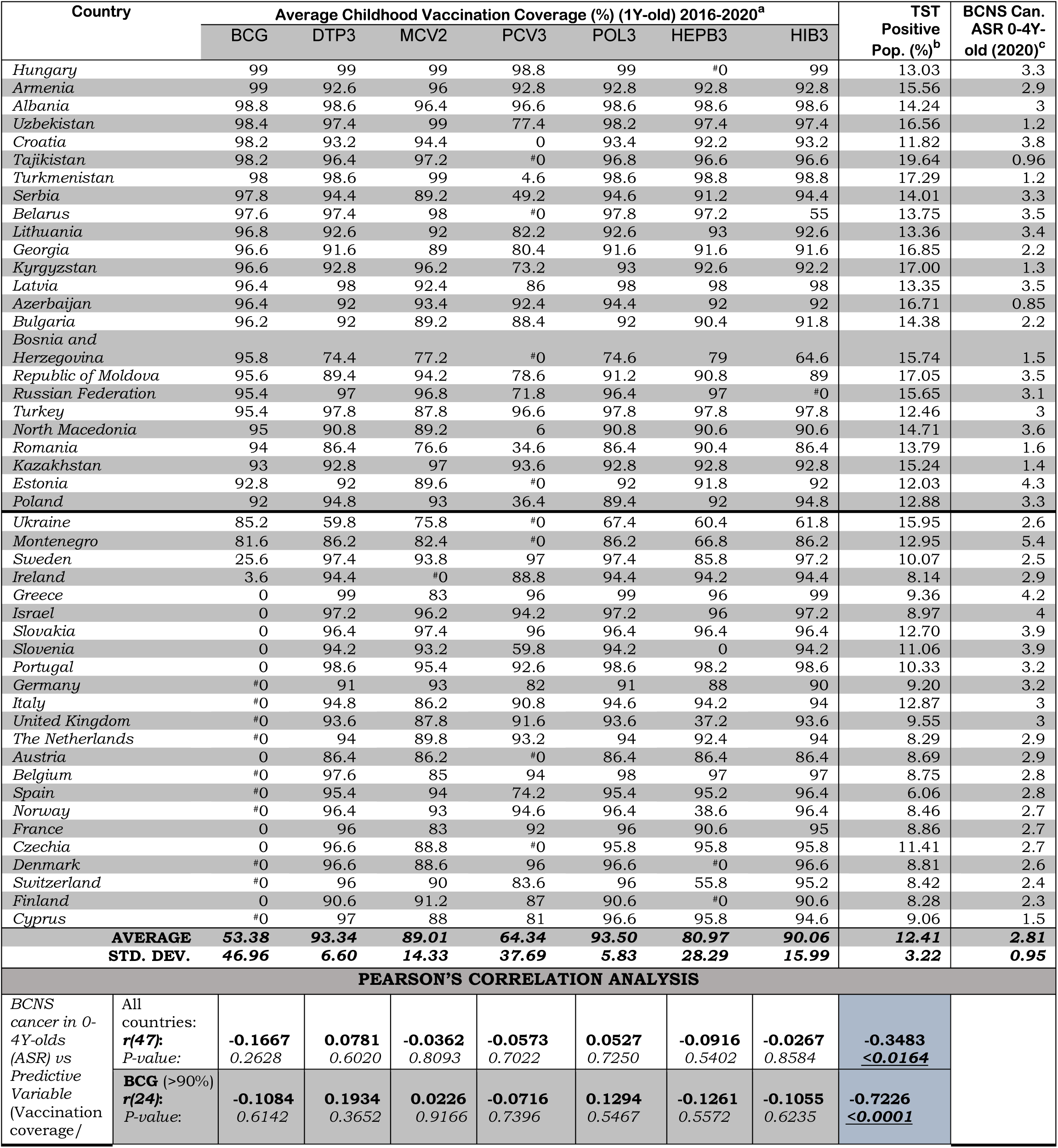
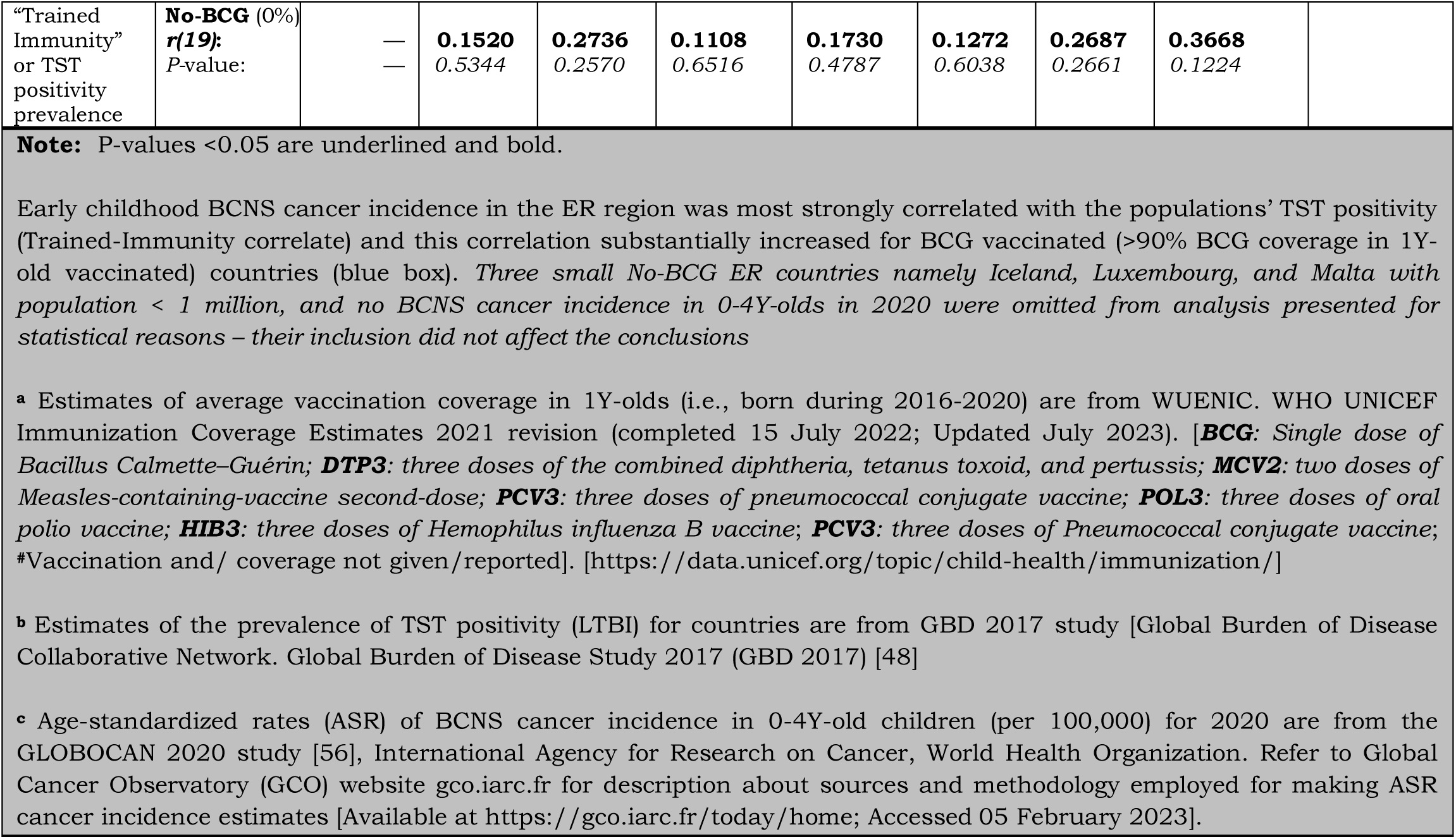
BCNS Cancer incidence in 0-4Y-old (<5Y-old) children in European Region: Vaccination coverage in <1Y-old, ‘Trained-Immunity’ prevalence from *Mycobacterium spp*. exposure) of populations (TST Positivity) and correlation analysis.

### Early childhood BCNS cancer incidences in ER countries are negatively associated with the prevailing possibility of *Mycobacterium* spp. exposure

Tuberculin immunoreactivity (TI), as measured by TST and interferon gamma release assays (IGRAs), is employed to indirectly measure the elicited or persisting cell-mediated immune response resulting from exposure to *Mycobacterium* spp. (BCG or environmental) antigens, as well as to ascertain BCG vaccine efficacy [49,50,57]. In the absence of a clinically active tuberculosis disease, it is also referred to as LTBI by the WHO for tuberculosis management purposes, due to these individuals supposedly “being at risk of developing TB” in their lifetime from reactivation or fresh infections [49,50,51,59]. Intriguingly, the prevalence of TI/LTBI in the European Region countries [48], which provides a measure of the cell-mediated trained-immunity persistence in the populations resulting from their exposure to *Mycobacterium* spp., including that from the BCG vaccine, was found to be significantly negatively correlated with BCNS cancer incidence rates in 0-4Y-olds [Pearson’s correlation coefficient, *r*(47): -0.3483, *p*-value: <0.0164; see Table 1 and Figure 1A]. If there is any protective role for *Mycobacterium* spp. exposure in reducing the incidence rate of early childhood BCNS cancer, it may be conjectured that the TI/LTBI prevalence would be more strongly but negatively correlated with early childhood BCNS cancer incidence in countries with neonatal BCG vaccination in place than in countries without BCG vaccination (no-BCG). Surprisingly, the negative correlation between BCNS cancer incidence rates and prevailing TI substantially improved for BCG-vaccinated countries (n=24; coverage >90%) [*r*(24): -0.7226, *p*-value: 0.0001], whereas for non-BCG-vaccinated or No-BCG countries (n=19), it became uncorrelated [*r*(19): 0.367, *p*-value: 0.122] (Table 1 and Figure 1B). A similar correlation was observed for the incidence of BCNS cancer in 2022 (data not shown). The country-wise BCG vaccination coverage in 0-4Y-olds in the European region is displayed on the map in Figure 1C. Note that BCG vaccination is common in the Eastern region (countries colored blue), while it is absent in Western regions (countries colored white). Furthermore, the inclusion of BCG countries [Ukraine, BCG coverage: 85.2%, BCNS cancer ASR: 2.6, TI/LTBI prevalence: 15.96%; Montenegro, BCG coverage: 81.6%, BCNS cancer ASR: 5.4, TI/LTBI prevalence: 12.96%] or No-BCG countries [Sweden, BCG coverage: 25.6%, BCNS cancer ASR: 2.5, TI/LTBI prevalence: 10.07%; Ireland, BCG coverage: 3.6%, BCNS cancer ASR: 2.9, TI/LTBI prevalence: 8.14%] that had BCG coverage other than >90% or 0% in the BCG and No-BCG group countries did not change the nature of the correlation. Thus, the observed strong negative correlation between BCNS cancer incidence in 0-4Y-olds of neonatal BCG-mandating countries and their prevailing immunoreactivity to mycobacterial antigens, but not in non-BCG-mandating countries (No-BCG), indicates a potential protective association that needs further investigation. Overall, based on the existing incidences, it may be argued that the combination of neonatal BCG vaccination priming of the developing immune system of neonates and its subsequent boosting on re-exposures to *Mycobacterium* spp. (BCG or environmental) may be protective in early childhood BCNS cancer incidences.

**Figure 1.**
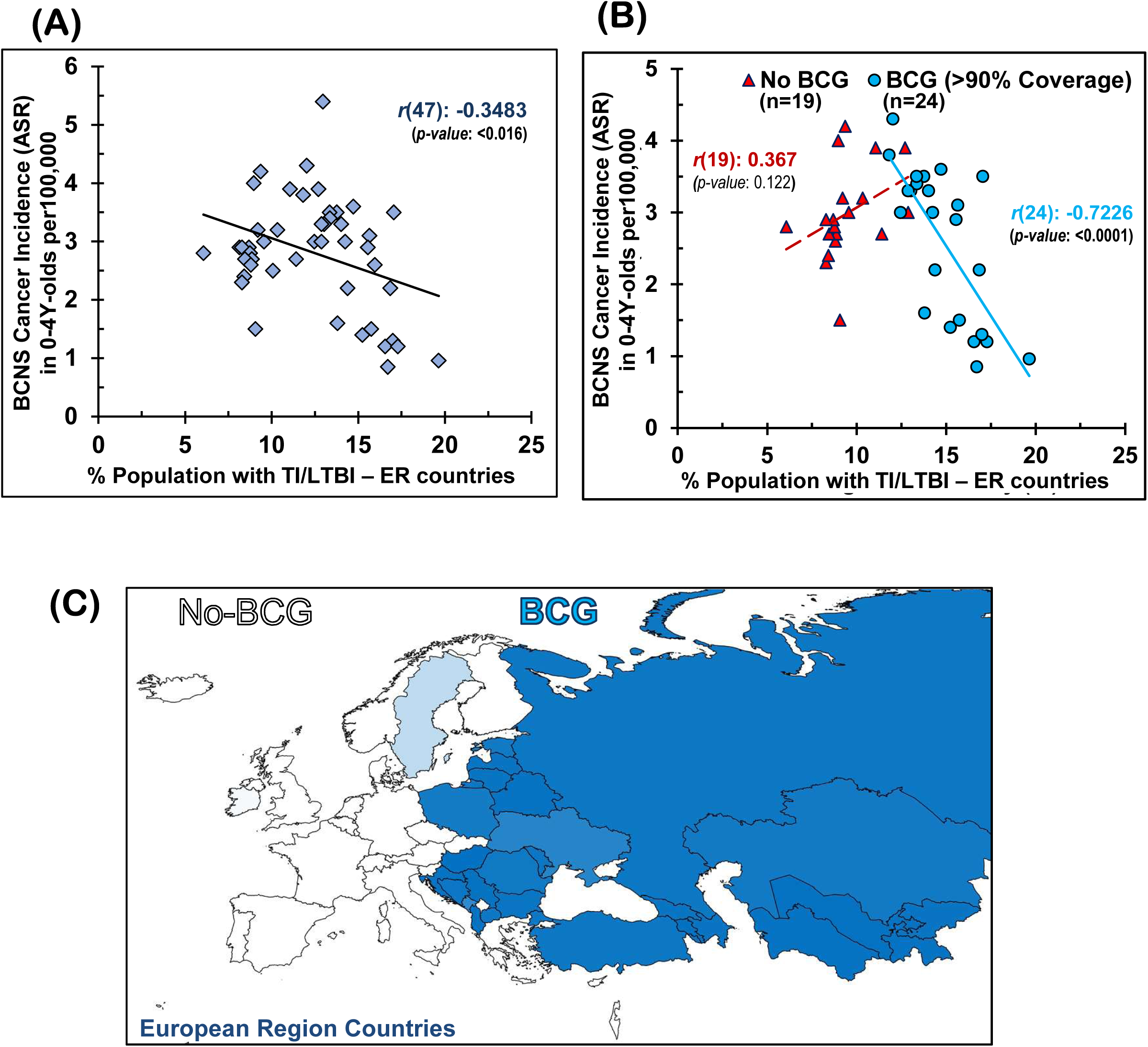
**Early childhood BCNS cancer incidences in neonatal BCG vaccination-implementing European Region (ER) countries were negatively correlated with their prevailing tuberculin immunoreactivity (TI) or LTBI estimates, (A)** without regard for BCG vaccination mandate and coverage, and **(B)** with regard to effective BCG vaccination (BCG) or lack thereof (No-BCG). (For correlation with other vaccines (e.g., BCG, DTP3, MCV2, PCV3, POL3, HIB3, HEPB) and their coverage refer to *Supplementary Figure. 1*). **(C)** BCG coverage in 0-4Y-olds in ER-countries. *BCG: Bacille Calmette-Guérin; BCNS Cancer: Brain and Central Nervous System Cancers; LTBI: Latent Tuberculosis infection*

Furthermore, this association of early childhood BCNS cancer incidence rate with the TI prevalence of countries was found to be more strongly correlated in the group of countries with effective BCG coverage (>90%) than any other early childhood vaccine coverage for country subgroups, as may be supposed from their coefficients of determination (R^2^ values), i.e., BCG (n=24): R^2^ = 0.5221; DTP3 (n=41): R^2^ = 0.1680; MCV2 (n=26): R^2^ = 0.2315; PCV3 (n=18): R^2^ = 0.209; POL3 (n=41): R^2^ = 0.1386; HEPB3 (n=34): R^2^ = 0.2052; and HIB3 (n=39): R^2^ = 0.1954 [see data points shown by blue and orange circles in Figure 2 a–g]. For vaccine coverage lower than 90%, most correlations are even weaker and statistically insignificant (p-value: >0.05), [see data points shown by red triangles in Figure 2 a–g]. Additionally, exploratory multiple regression analysis indicated that the coverage of other vaccines (co-vaccination) or their combinations also does not seem to affect BCNS cancer incidence reduction in less than 5-year-old children, in addition to TI in the neonatally BCG-vaccinated populations. Interestingly, when all countries are considered, completely disregarding their neonatal BCG vaccination policy and coverage, the countries with medium-to-high BCNS cancer incidence rates (i.e., ASR 2–4 and >4 per 100,000) exclusively belong to low TI/TST-positive populations (<15%), whereas the low BCNS cancer incidence countries (i.e., ASR >0– 2 per 100,000) are primarily BCG-vaccinating countries (8/9) with high prevailing TI/TST-positive populations (7/9 with >15–20%). Overall, early childhood BCNS cancer incidence in countries that use neonatal BCG vaccination is strongly but negatively linked to TI/TST positivity in the community. These findings suggest that neonatal BCG vaccination plays a protective role in countries where the immune system of any child is more likely to be boosted by exposure to environmental *Mycobacterium* spp. Together, these results imply that environmental *Mycobacterium* spp. exposure-associated boosting in children primed with neonatal BCG vaccination may play an important role in lowering the incidence of BCNS cancer in early childhood (less than 5 years of age).

**Figure 2.**
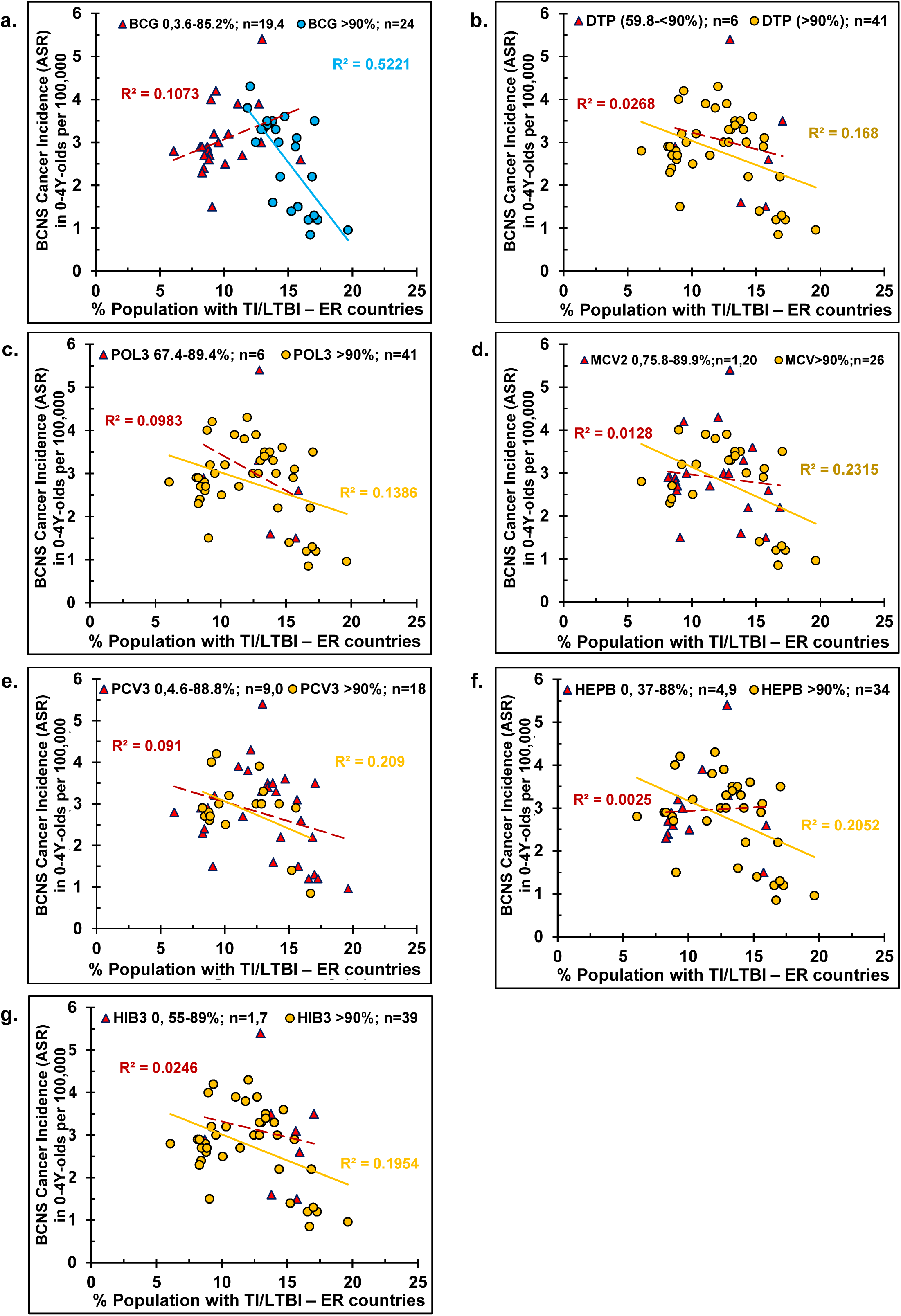
The negative correlation of BCNS cancer incidence in 0-4Y-old with Tuberculin Immunoreactivity (TI/LTBI/TST-positivity) of the population (likelihood of *Mycobacterium* spp. exposure) was most strongly associated with the priming of the majority of children(>90%) by neonatal BCG vaccination **(a),** but not with the coverage of any other commonly employed childhood vaccine, i.e., **(b)** DTP3, **(c)** POL3, **(d)** MCV, **(e)**PCV3, **(f)** HEP3, **(g)** HIB3 [n= number of countries; R^2^: coefficient of determination; solid and broken trend line are for *p*-value <0.05 and >0.05, respectively]

## DISCUSSION

Regarding the impact of potential confounders on our observations and outcomes of any prospective study, it must be remembered that many previous studies have positively associated high BCNS incidence rates with socioeconomic position (SEP), non-chromosomal structural defects, a high birth rate, syndromes (*e.g*., neurofibromatosis types I and II, Li Fraumeni syndrome, tuberous sclerosis), and polymorphisms in a number of genes and infections [reviewed in 11,12]. Infection of *Toxoplasma gondii*, a protozoan, has also been associated with an increased risk of glioma (a subtype of BCNS cancers). Human leukocyte antigen (HLA) alleles, composition of immune cells, and the genomic architecture of T-, NK-, and myeloid cells have been associated with glioma risk. However, the role of most viral infections studied has been inconsistent in glioma, a malignant BCNS cancer that is responsible for the majority of deaths. Only allergies, atopic conditions, and infection with Varicella Zoster Virus (VZV), a herpes virus that causes chickenpox and shingles, have been consistently associated with a reduced risk of glioma. The existence of a potential protective association between environmental *Mycobacterium tuberculosis* complex spp. exposure and BCNS cancer incidence has been suggested by us based on the observed lower incidence rates in high tuberculosis reporting countries [45]. These may very well remain underlying confounders for swaying the outcome of any exploratory study and hence the conclusions, including those of the current article. Their control would be desirable in future studies.

No matter how significant and strong any statistical correlation could appear between potential variables, it can never be considered to have a cause-and-effect relationship. Endeavors may be made to identify and validate protective variables that have a potential cause-and-effect relationship [51]. The channelization of resources to evaluate and explore the preventive and protective potential of childhood vaccinations, especially BCG vaccination and boosting events (BCG and environmental *Mycobacterium* spp.), may be warranted to ascertain a potential cause-and-effect relationship. It should also be remembered that there are certain inherent lineage-specific and preparation-specific differences with regard to content and immunogenicity in BCG sub-strains [60,61] that are historically differentially employed in the Eastern and Western European region countries for children’s vaccination [47], as well as differences in circulating *Mycobacterium* spp. [62-64] and their inherent immune activation potential [65-67] on exposure. Co-vaccination during this period may also have an impact [27,68]. While designing and performing any such exploratory study, more emphasis should be placed on identifying variables potentially causatively associated with direct reduction in early childhood BCNS cancer incidence (having cause-and-effect relationships) with some interventional value rather than just being an explanatory variable (e.g., income, GDP, socioeconomic position in society, gene polymorphism number of children, etc. [11,12]). Additional study design considerations may be needed to control the effect of possible confounders that have been previously associated with BCNS cancer incidences and could be supposedly overrepresented in any specific study population (e.g., syndromes, gene polymorphisms, mutations in genes associated with immune system functioning, etc.; see the previous paragraph). The possible modulation of the developing immune systems of children resulting from their interactions with their mother and other closely interacting individuals or social contacts, the potential reduction of the transfer of unknown causatively associated etiological agents to young children in a high TI population, or the potential of endocrine or *in utero* reprogramming by mothers exposed to mycobacterial antigens or other pathogens may also be considered as possible variables for any clinical trial attempting to evaluate the protective potential of the BCG vaccine, as previously suggested for leukemia [37,38,43].

## CONCLUSION

Exposure to *Mycobacterium* spp. (both BCG and environmental) may potentially contribute to a decrease in BCNS cancer incidence during early childhood in countries that follow neonatal BCG vaccination. Dedicated epidemiological studies exploring links between BCNS cancer incidence during early childhood (less than 5 years old) and childhood vaccinations, pathogen exposures, and immune training are warranted, utilizing associated children’s health records. It should be backed by follow-up randomized controlled clinical trials, preferably performed in populations with low prevalence of tuberculin immunoreactivity (TI/LTBI), explicitly exploring the impact of BCG vaccination and its boosters on early childhood BCNS cancer incidence while suitably controlling for the underlying heterologous cell-mediated immunity and “trained immunity” correlates and other possible confounders as indicated in the preceding paragraphs to conclusively determine the biological significance of the observed association and its potential practical/interventional application, if any.

## MATERIALS AND METHODS

### Data sources

The BCNS cancer incidence in different European region countries’ children (both male and female) who are less than 5 years of age (0-4Y-old) in 2020 was obtained from the ‘Global Cancer Observatory (GCO), WHO GLOBOCAN 2020, The International Agency for Research on Cancer’ (IARC), WHO [56] without applying any additional exclusion criteria. The relevant years’ early childhood vaccination coverage data was from ‘Estimates of National Immunization Coverage (WUENIC): 2022 revision, updated July 2023, WHO-UNICEF’ [46]. The prevailing tuberculin immunoreactivity (TI) or TST positivity data for the countries’ population was from latent tuberculosis infection (LTBI) estimates of the Global Burden of Disease Collaborative Network. Global Burden of Disease Study 2017 (GBD 2017) Results. Seattle, United States: Institute for Health Metrics and Evaluation (IHME), 2018 [48].

### Inclusion and Exclusion criteria

The data for WHO’s European region countries were extracted from the above-indicated sources for analysis. The data on cancer incidence, vaccine coverage, and prevailing LTBI/TST was an aggregate for the countries without any segregation by race, sex, or residency of parents. Countries reporting no incidence of BCNS cancer in 0-4Y-olds (i.e., Iceland, Luxembourg, Malta) were excluded from association analysis. No additional inclusion or exclusion criteria were employed.

### Statistical Analysis and Figure Generation

The exploratory associative analysis was performed on the European Region countries’ data using routine statistical methodology in Excel 2019, as described previously [51,69]. Pearson’s correlation coefficient determination and regression analysis were performed. Simple and multiple regressions were performed with different predictor variables (e.g., coverage of different vaccines and tuberculin reactivity/TST/LTBI prevalence). Countries with >90% vaccine coverage and lower were grouped separately for analysis as indicated in the results. Pearson’s correlation coefficient (*r*) value of -0.6 to -0.8 was considered a strongly negative correlation. Statistically, the p-value of <0.05 was considered significant. Diagrams were generated in Excel 2019, and the choropleth map displaying differential BCG coverage across ER was generated using Inkscape 1.0. Randomization, blinding, and power analysis were not applicable for the study. No new codes were generated.

## DATA AVAILABILITY STATEMENT

The primary data used for the analysis and included in the manuscript are available at the above indicated sources. Data generated by the authors are included in the article.

## FUNDING STATEMENT

The current study did not receive any funding from any source.

## Data Availability

All data produced in the present work are contained in the manuscript.

## ACKNOWLEDGEMENTS

The general funding support from Banaras Hindu University (IoE-BHU and IMS) to the laboratory of SS is acknowledged. The funders had no role in the study design, data collection and analysis, decision to publish, or manuscript preparation.

## ETHICAL STATEMENT

The study complied with the existing ethical standards.

## COMPETING INTERESTS

The authors have declared that no competing interests exist.

## Notes

### Competing Interest Statement

The authors have declared no competing interest.

### Funding Statement

This study did not receive any funding

### Author Declarations

The study used (or will use) ONLY openly available human data that were originally found atGCO. WHO GLOBOCAN 2020 (2023). Global Cancer Observatory, The International Agency for Research on Cancer (IARC), WHO. CEDEX, France [Available/located at: https://gco.iarc.fr/today/home]

